# Persisting decreases in state and trait anxiety post-psilocybin: A naturalistic, observational study among retreat attendees

**DOI:** 10.1101/2022.03.02.22271743

**Authors:** M.K. Kiraga, K. P. C. Kuypers, M. V. Uthaug, J.G. Ramaekers, N. L. Mason

## Abstract

Anxiety disorders are the most common type of psychiatric disorders among Western countries. Evidence-based treatment modalities including pharmacological and cognitive-behavioral therapy result in relatively low response rates (average range: 51 - 58%). Historical and recent research suggests psychedelic drugs may be efficacious in alleviating anxiety-related symptoms among healthy and clinical populations. The main aim of the present study was investigation of the effects of psilocybin-containing truffles, when taken in a supportive group setting, on ratings of state and trait anxiety across self-reported healthy volunteers. Attendees of psilocybin ceremonies were asked to complete a test battery at three separate occasions: before the ceremony (baseline), the morning after, and one week after the ceremony. The test battery included questionnaires assessing state and trait anxiety (State-Trait Anxiety Inventory), mindfulness capacities (Five Facet Mindfulness Questionnaire), and personality (Big Five Inventory). Additionally, the psychedelic experience was quantified with the Persisting Effects Questionnaire and the Ego Dissolution Inventory. The total amount of psilocybin-containing truffles consumed by each participant was recorded, and a sample of the truffles was analyzed to determine psilocin concentrations. Fifty-two attendees (males= 25; females= 25; others= 2) completed parts of the baseline assessment, 46 (males= 21; females= 24; others= 1) completed assessments the morning after the ceremony, and 23 (males= 10; females= 13) completed assessments at the one-week follow-up. Average psilocin consumption across individuals was 27.1 mg. We observed medium to large reductions in anxiety measures (both state and trait) compared to baseline which persisted over a one-week period post-ceremony. At one week post-ceremony, the non-judging facet of the mindfulness scale was increased, while the personality trait neuroticism decreased, when compared to baseline. Additionally, we found neuroticism and ratings of ego dissolution to be the strongest predictors of reductions in trait and state anxiety, respectively. In sum, results indicate rapid and persisting (up to one week) anxiolytic effects in psilocybin retreat attendees, which are related to the acute experience of ego dissolution, as well as lasting changes in trait neuroticism. To understand whether these effects extend to wider populations suffering from heightened anxiety, and the mechanisms involved, further experimental research is needed.

## 1. Introduction

Anxiety disorders (e.g., generalized anxiety disorder, panic disorder, social phobia) are the most common psychiatric disorders, with a current worldwide prevalence of 7.3% (Sansone and Sansone 2010; Stein et al. 2017). They were also found to be burdensome and costly to individuals and wider society (Kandola et al. 2018). Despite such a widespread presence, current treatments only provide partial success rates, with pharmacological therapies resulting in response rates of 50.8%-58.3% (Baldwin 2008; Otto et al. 2010), and cognitive behavioral therapies (CBT) resulting in response rates of 51.7%-53.6% (Loerinc et al. 2015; Otto et al. 2010) in adults with anxiety disorders. Additionally, the combination of the two approaches has not been found to result in more promising outcomes, with a response rate of 54.2% (Otto et al. 2010). Further studies have concluded that the exact rate of treatment response also varies per anxiety disorder. For instance, in patients with a lifetime panic disorder, the response rates drop drastically to about 13% for each of the aforementioned therapeutic modalities (Brown et al. 1996), whereas patients with comorbid anxiety and depression tend to prematurely terminate treatment more frequently than patients with a single diagnosis (Brown et al. 2001). The above findings illustrate a rising urgency within the field of mental health for efficient fast-acting treatment options that prove efficacious for the large heterogeneous cluster that makes up anxiety disorders, and that persist after treatment.

Before criminalization of psychedelic drugs in the 1970s, studies combining administration of a psychedelic drug and psychological therapy found significant improvements in anxiety-related symptomatology (Weston et al. 2020). Recently, there has been a resurgence of interest regarding the therapeutic potential of these substances, including psilocybin, ayahuasca, and LSD, in treating an array of different mental health disorders (Carhart-Harris et al. 2016; Grob et al. 2011; Osório et al. 2015; Palhano-Fontes et al. 2019; Sanches et al. 2016). More modern trials have assessed the efficacy of psychedelics to reduce anxiety symptoms among patients with a life-threatening illness, and have demonstrated large significant reductions in ratings of anxiety, with a response rate of 60-80% sustained over the course of 6 to 12 months post treatment (Gasser et al. 2014; Griffiths et al. 2016; Ross et al. 2016), and in a smaller cohort up to 4.5 years post treatment (Agin-Liebes et al. 2020). Similar reductions in ratings of anxiety symptoms have been demonstrated in trials assessing the efficacy of a psychedelic to alleviate symptoms of treatment-resistant depression (Carhart-Harris et al. 2018; Carhart-Harris et al. 2016; Osório et al. 2015), and in naturalistic samples with heterogeneous mental health status (Jiménez-Garrido et al. 2020; Uthaug et al. 2019; Uthaug et al. 2018). In healthy volunteers, the persisting effect of psychedelics on feelings of anxiety have been mixed, with one study reporting persisting reductions in acute (state) anxiety one week, and reductions in personality-related (trait) anxiety one month after psilocybin ingestion (Barrett et al. 2020), another study reporting no changes in trait anxiety 1 and 12 months after LSD (Schmid and Liechti 2018). The questionable rigidity in the research design of the earlier studies (Weston et al. 2020) and the promising findings of modern trials warrant further investigation of the persisting effects of ingestion of a psychedelic drug on acute and personality-related anxiety, independent of life-threatening illness or comorbid depression. Furthermore, if psychedelic substances *do* reduce anxiety, it is of interest to understand what psychological processes may be at play.

The personality trait neuroticism, reflecting an individual’s tendency to experience negative emotions, sensitivity to aversive cues, and insufficient ability to cope with stress, has been found to be the personality trait that is the most strongly associated with many forms of psychopathology (Caspi et al. 2005; Clark 2005; Frokjaer et al. 2010; Klein et al. 2011; Krueger and Tackett 2003; Ormel et al. 2013), and a core vulnerability factor common to anxiety and mood disorders (Zinbarg et al. 2016). Although it has been argued that personality traits are relatively stable throughout adulthood, an array of clinical work has shown that personality is amenable to changes across the lifespan (Roberts et al. 2017), with neuroticism being found to fluctuate to one standard deviation across the lifespan (Roberts 2006). As such, it has been suggested that reducing neuroticism should be the primary focus of therapeutic interventions when treating psychopathologies such as anxiety (Roberts et al. 2017). It has been repeatedly found that ingestion of a psychedelic drug can induce rapid and persisting changes in personality traits, and particularly reductions in neuroticism, in both healthy and clinical populations (Barrett et al. 2020; Bouso et al. 2012; Bouso et al. 2015; Erritzoe et al. 2018; Johnstad 2021; Kiraga et al. 2021; Nour et al. 2017; Weiss et al. 2021). Thus, given the reduction in self-rated neuroticism found after ingestion of a psychedelic, it could be suggested that one mechanism by which psychedelic substances induce persisting changes in anxiety levels could be by altering maladaptive personality structures which exacerbate anxiety. However, the relationship between psychedelic-induced changes in anxiety and psychedelic-induced changes in neuroticism has yet to be assessed.

A further psychological processes which may be of particular salience when discussing therapeutic efficacy for anxiety, and which psychedelics have been repeatedly found to enhance, are mindfulness capacities (Carhart-Harris et al. 2016; Eleftheriou and Thomas 2021; Madsen et al. 2020; Murphy-Beiner and Soar 2020; Payne et al. 2021; Uthaug et al. 2020; Uthaug et al. 2019; Uthaug et al. 2018). Mindfulness has been described as “paying attention in a particular way: on purpose, in the present moment, and non-judgmentally” (Kabat-Zinn 2009). It is generally considered multifaceted, including capacities such as being able to: notice or attend to internal feelings, thoughts, and external simulation (‘observing’), label feelings, thoughts and experience with words (‘describing’), attending to what is happening in the present moment (‘acting with awareness’), take a non-evaluative stance toward internal thoughts and feelings (‘non-judgement of inner experience’), and being able to allow emotions and thoughts to come and go, without being interfered by them (‘non-reactivity to inner experience’). Given that one of the core components of anxiety disorders is the practice of excessive rumination on non-present fears, leading to a vicious circle of higher levels of physical arousal and more worrying (Dar and Iqbal 2015), individuals suffering from anxiety disorders typically lack or have lower levels of mindfulness capacities (Goldin and Gross 2010). Accordingly, it has been shown that interventions which enhance mindfulness capacities, such as mindfulness-based stress reduction training (MBSR), can be an effective form of treatment for people with anxiety disorders (Ramel1 et al. 2004), resulting in higher efficiency and improvement rates than cognitive-behavioral group therapy (Koszycki et al. 2007). That said, the relationship between psychedelic-induced changes in mindfulness capacities, and psychedelic-induced changes in anxiety, has yet to be assessed.

Finally, another important factor when thinking about therapeutic applications is the longevity of effects. Usually when taking conventional anxiolytics the treatment duration will vary but might extend over months (Loerinc et al. 2015; Otto et al. 2010). For psychedelics, persisting effects on personality or wellbeing can be found up to 12 months after a single dose administration (Schmid and Liechti 2018), whereas increased mindfulness capacities have been found to persist up to 3 months (Madsen et al. 2020). It has been suggested by some studies that the intensity or quality of the psychedelic experience determines the treatment outcome (Griffiths et al. 2018; MacLean et al. 2011; Roseman et al. 2018). Specifically, it has been repeatedly found that psychedelic-induced mystical-type experiences and ego dissolution, the latter a phenomenon characterized by the reduction in self-referential awareness, disruption self-world boundaries, and increased feelings of unity with one’s surroundings (Nour and Carhart-Harris 2017), correlates with long-term (positive) outcomes (Bogenschutz et al. 2015; Griffiths et al. 2011; Griffiths et al. 2018; Garcia-Romeu et al. 2014; MacLean et al. 2011; Roseman et al. 2018; Uthaug et al. 2018). Thus, in the present study we also assessed whether we could predict persisting changes in anxiety based on acute ratings of ego dissolution.

In sum, anxiety disorders evoke a notable impact on people’s lives, yet the existing treatment options are of limited success. Based on this and previous findings of psychedelic-induced alleviation of anxiety, the present study was designed to assess the sub-acute effects of psilocybin in retreat attendees on state and trait anxiety, when taken in a supportive group setting. The latter point is highlighted, as previous research has suggested that the combination of psychedelic drug administration with (psychological) support is necessary to evoke a (treatment) response (Weston et al. 2020). Additionally, we aimed to replicate the previously reported findings on the substance’s ability to increase mindfulness capacities and decrease neuroticism (Carhart-Harris et al. 2016; Erritzoe et al. 2018; Johnstad 2021; Kiraga et al. 2021; Madsen et al. 2020; Murphy-Beiner and Soar 2020; Uthaug et al. 2020; Uthaug et al. 2019; Uthaug et al. 2018). As previously rationalized, that such changes may be related to reductions in anxiety, we further aimed to assess whether changes in mindfulness capacities and neuroticism correlated with changes in anxiety. Lastly, we wanted to assess the relationship between psychedelic-induced changes in state and trait anxiety and ego dissolution.

We hypothesized that, compared to baseline, reductions in state and trait anxiety and neuroticism would be observed 24 hours and 7 days after ingestion of psilocybin, whereas mindfulness capacities would be enhanced. Furthermore, we hypothesized that reductions in ratings on both aspects of anxiety would be negatively correlated with enhancements in mindfulness capacities and positively correlated with reductions in the personality trait, neuroticism. Finally, we hypothesized that positive changes in ratings of anxiety would be correlated with higher subjective ratings of ego dissolution, the latter measured with the Ego Dissolution Inventory (Nour and Carhart-Harris 2017).

## 2. Methods

### 2.1 Participants and study procedure

Participants were volunteers attending legal psilocybin retreats in the Netherlands, organized by the Psychedelic Society UK. Attendees of those ceremonies were either invited to participate in the study on site, or contacted the researchers by email after hearing about the study through the retreat organizers. To participate in the study, volunteers had to be a minimum of 18 years old and proficient in English. Participants were assessed 3 times: at baseline (the evening before psilocybin ingestion), within 24 hours after psilocybin ingestion (hereafter referred to as: sub-acute) and 7 days after psilocybin ingestion (hereafter referred to as: follow-up). Seven-days later, participants received the final follow-up measurement online (through Qualtrics). The total amount of truffles (g) taken by each participant was recorded, and a sample of the truffles was taken to determine the concentrations of alkaloids afterwards.

The study was conducted in accordance with the Declaration of Helsinki and subsequent amendments concerning research in humans and was approved by the Ethics Review Committee of Psychology and Neuroscience and Maastricht University (ERCPN-175_03_2017_A5). Participation was voluntary and no incentives to participate were provided. All volunteers gave their written informed consent prior to participation. The research team was not involved in the screening, preparation, organization, administration, and supervision of the psilocybin ceremonies that were visited.

### 2.2 Psilocybin retreats

Prior to participation in the retreat, personal intakes were done by the facilitators, which included screening for (and excluding) individuals with psychiatric disorders or taking psychiatric medications, and medical factors like high blood pressure.

The setting in which psilocybin was taken was the same throughout all of the retreats. Participants stayed in a large house set in nature, hosted by at least two or more experienced psilocybin facilitators. They arrived the evening before psilocybin administration, and were able to get acquainted with each other, the facilitators, and the schedule of the retreat. The next day, participants received the psilocybin-containing truffles around noon, in a tea form. Facilitators provided music, tools to draw and/or write, food, and overall support. In the evening, all participants and facilitators came back together as a group. The next morning, all participants had breakfast together and had a closing group meeting.

### 2.3 Psilocybin

Participants ingested the truffles in a tea form, guided by the facilitators. To do this, the truffles were crushed, and boiling hot ginger tea was added. After infusing for a few minutes, the participants drank the tea, and were subsequently free to add more water and repeat the process 2–3 times. Afterwards participants could eat the remaining truffle contents in the cup. Previous experimental studies have demonstrated that subjective alterations after psilocybin intake begin 20–40 minutes following administration, peak around 60–90 minutes, and subside by six hours post-intake (Hasler et al. 2004).

However, anecdotal reports suggest that when ingested in tea form, subjective alterations are felt more quickly, and for a shorter amount of time (Erowid 2015).

The total amount of psilocybin truffles taken by each participant was recorded, and a sample of the truffles was taken to determine concentrations of psilocybin and its metabolite, psilocin. The German Central Customs Authority determined the contents of psilocin and psilocybin after freeze-drying the truffles using a previously described HPLC method (Laussmann and Meier-Giebing 2010).

### 2.4 Assessments

The assessments included a basic demographic section and six questionnaires: the State-Trait Anxiety Inventory (STAI), the Five Facet Mindfulness Questionnaire (FFMQ), the Big Five Inventory (BFI), the Persisting Effects Questionnaire (PEQ), and the Ego Dissolution Inventory (EDI). All materials were provided in English. The STAI and FFMQ were filled out three times, i.e., at baseline, sub-acute session, and follow-up. The BFI was administered twice, during the baseline and the follow-up session. Whereas the remaining 2 questionnaires were only completed once; the EDI during the sub-acute session, to assess the magnitude of the psychedelic experience in retrospect, and the PEQ at the 7 day follow-up, to assess persisting effects and the significance of the experience.

#### 2.4.1 State-Trait Anxiety Inventory (STAI)

The State-Trait Anxiety Inventory (STAI; Spielberger 1983) is a 40-item rating scale with a 4-point response format, ranging from 1 (*almost never*) to 4 (*almost always*) that is scored into two sub-scales (state anxiety and trait anxiety). For “state” anxiety questions, participants were asked to select the response for each item that best describes how they feel “right now, that is, at this moment”. For “trait” anxiety questions, participants were asked to select the response that best describes how they “generally feel, that is, most of the time”. The reverse-scored and (e.g., “I feel pleasant”) and direct-scored (e.g., “I feel nervous and restless”) items are summed per each subscale to create a total score which ranges between 20 and 80, with higher scores indicating greater anxiety. Internal consistency coefficients for the scale have been shown to range from .86 to .95 and test-retest reliability coefficients from .65 to .75 over a 2-month interval (Spielberger 1989). Considerable evidence attests to the construct and concurrent validity of the scale (Spielberger 1989).

#### 2.4.2 Five Facet Mindfulness Questionnaire (FFMQ)

The Five Facet Mindfulness Questionnaire, 39 items,(FFMQ; Baer et al. 2006) measures five different factors: 1) Observe: noticing external and internal experiences, e.g., body sensations, thoughts, or emotions; 2) Describe: putting words to, or labeling the internal experience; 3) Acting with awareness: focusing on the present activity instead of behaving mechanically; 4) Non-judging the inner experience: taking a non-evaluative stance towards the present experience, thoughts, or emotions; and 5) Non-reacting to the inner experience: allowing thoughts and feelings to come, without getting caught up in, or carried away, by them. Sample item of the Observe dimension is: “When I take a shower or bath, I stay alert to the sensations of water on my body”. Participants were asked to rate the degree of concordance with each statement on a 5-point Likert scale that ranges from 1 (*never or very true*) to 5 (*very often or always true*), for 39 statements. The FFMQ has shown adequate psychometric properties in both non-clinical and clinical samples. Cronbach’s α for each individual sub-scale range from 0.75 to 0.91 (Baer et al. 2006), and the internal consistency of the scale in our sample was of 0.83 and 0.86 (pre- and post-intake, respectively).

#### 2.4.3 Big Five Inventory (BFI)

The Big Five Inventory (BFI-44; John, Donahue and Kentle 1991) was used to measure the Big Five personality dimensions, specifically: 1) Extraversion (8 items); 2) Agreeableness (9 items); 3) Conscientiousness (9 items); 4) Neuroticism (8 items) and Openness (10 items), total of 44 items. Those prototypical traits defining each of the Big Five dimensions are assessed by short and easy-to-understand phrases, for example: “I see myself as someone who is talkative”. The items are rated on a 5-point Likert scale ranging from 1 (*disagree strongly)* to 5 (*agree strongly*). The direct and reverse-scored items for each dimension are summed together to create a total score, which, given the variability in terms of number of items, ranges between 8-40, 9-45, or 10-50. The BFI scales have shown substantial internal consistency, retest reliability, and clear factor structure, as well as considerable convergent and discriminant validity with longer Big Five measures (Benet-Martínez and John 1998; John and Srivastava 1999).

#### 2.4.4 Persisting Effects Questionnaire (PEQ)

The Persisting Effects Questionnaire (PEQ) is a 143-item long scale aiming to assess changes in attitudes, moods, behavior, and spiritual experience (Griffiths et al. 2006). Prior research found that PEQ is sensitive to the prolonged effects of psychedelics, occurring even a one year after the ingestion (Schmid and Liechti 2018). Due to time constraints, the current study used a shortened version of the scale (90 items), including five out of six main categories: *attitudes about life* (Number of items (N)= 26); *attitudes about self* (N= 22); *mood changes* (N= 18); *relationships* (N= 18); and *behavioral changes* (N= 2). The 86 items of these five categories were rated on a 6-point scale (ranging from 0= *none* to 5= *extreme*). The scores of the resulting 10 scales (positive and negative scales for each of 5 categories) were assessed by calculating mean (SE) separately for each category.

The questionnaire also included four questions rated on a eight-point scale (1= *no more than routine*, and 8= *the single most meaningful experience of my life*): 1) “How personally meaningful was the experience?”; 2) “Indicate the degree to which the experience was spiritually significant to you?”; 3) “How psychologically challenging were the most psychologically challenging portions of the experiences?”; 4) “How personally psychologically insightful to you were the experiences?” (Griffiths et al. 2011; Johnson et al. 2014).

#### 2.4.5 Ego Dissolution Inventory (EDI)

The Ego Dissolution Inventory (EDI) is an eight-item self-report scale that assesses the participant’s experience of ego dissolution (Nour et al. 2016). In the present study, the original, English version was used to acquire a better understanding of the experiences the participants had about ego dissolution during the psilocybin ceremony. For example “I experienced a dissolution of my “self” or ego” and “I felt at one with the universe”. The participants answered the scale with endpoints of either 0 % (*No, not more than usual*) or 100 % (*Yes, I experienced this completely/entirely*). The EDI was scored by calculating the mean of all items, with a higher total score indicating a stronger experience of ego dissolution. The scale has been shown to have excellent internal consistency (Nour et al. 2016). The investigation of convergent validity of the scale has shown strong positive correlations between EDI and the mystical-type (or “peak”) of experience (Nour et al. 2016).

### 2.5 Statistical analyses

Data analyses and visualizations were performed using the Seaborn (version 0.11.2), Statsmodels (version 0.12.2), Scipy (version 1.7.1), and Patsy (version 0.5.1) packages for Python 3.8. A separate multi-linear regression model using the ordinary least square method (OLS) was fit for each outcome variable. The model included *Session* encoded as a dummy independent variable of three levels: baseline, the sub-acute and follow-up sessions. Subsequently, separate contrasts were performed between baseline and sub-acute as well as baseline and follow-up sessions.

Quantification of the acute experience was done by analyzing the results of EDI. Descriptive statistics (mean, standard deviation, range of scores) were calculated for the ratings of ego dissolution, together with a visual representation of kernel density estimation. To quantify the persisting functional outcomes of the experience, main outcome measures and additional questions of the PEQ were aggregated and reported in a tabular form.

Additionally, we sought to examine previously reported associations between anxiety and personality (Bienvenu et al. 2004; Cuijpers et al. 2005) as well as between anxiety and mindfulness capacities (Goldin and Gross 2010; Ramel1 et al. 2004). Given that ratings of ego dissolution have been shown to correlate with persisting effects after a psychedelic experience in both clinical and naturalistic studies (Bogenschutz et al. 2015; Griffiths et al. 2011; MacLean et al. 2011; Griffiths et al. 2018; Roseman et al. 2018; Uthaug et al. 2018), we tested for the possible associations between EDI scores and changes in anxiety. Canonical correlations (Sherry and Henson 2005) were conducted to evaluate the association between psilocybin-induced changes in (i) self-rated mindfulness capacities and neuroticism, (ii) ratings of ego dissolution, and (iii) state and trait anxiety. Variables were separated into two sets; set 1 included the psychological processes (i) and acute ratings of ego dissolution (ii) as predictors, and set 2 included the anxiety variables as criterion (iii). Canonical correlations were chosen as this approach assesses the relationship between two multivariate data sets, allowing investigation of variables that may have multiple causes and effects, while also reducing the potential of type 1 error (Sherry and Henson 2005).

For all statistical analyses, the alpha criterion level of statistical significance was set at p ≤ 0.05 and Cohen’s effect (d) size was reported in case of significant results to demonstrate the effect’s magnitude with 0.2-0.5 considered as small, 0.5-0.8 as a medium, and > 0.8 as large effect size (Leary, 2014).

## 3. Results

### 3.1 Participants and dose

Demographic information and concentrations of the psilocybin sample are all previously published elsewhere, and briefly summarized here (Mason et al. 2019).

#### Participants

Fifty-five volunteers agreed to participate in the present study and signed the informed consent. Of those 55 participants, 47% (N=26) identified themselves as males, 47% (N=26) as females, and 5% (N=3) reported identification with other gender categories. The exact numbers of participants’ enrollment and questionnaire completion are shown in Figure 1. Incomplete or missing test batteries were due to time constraints, as attendees must carry on with the semi-fixed retreat schedule, or participant drop-out.

**Figure 1.**
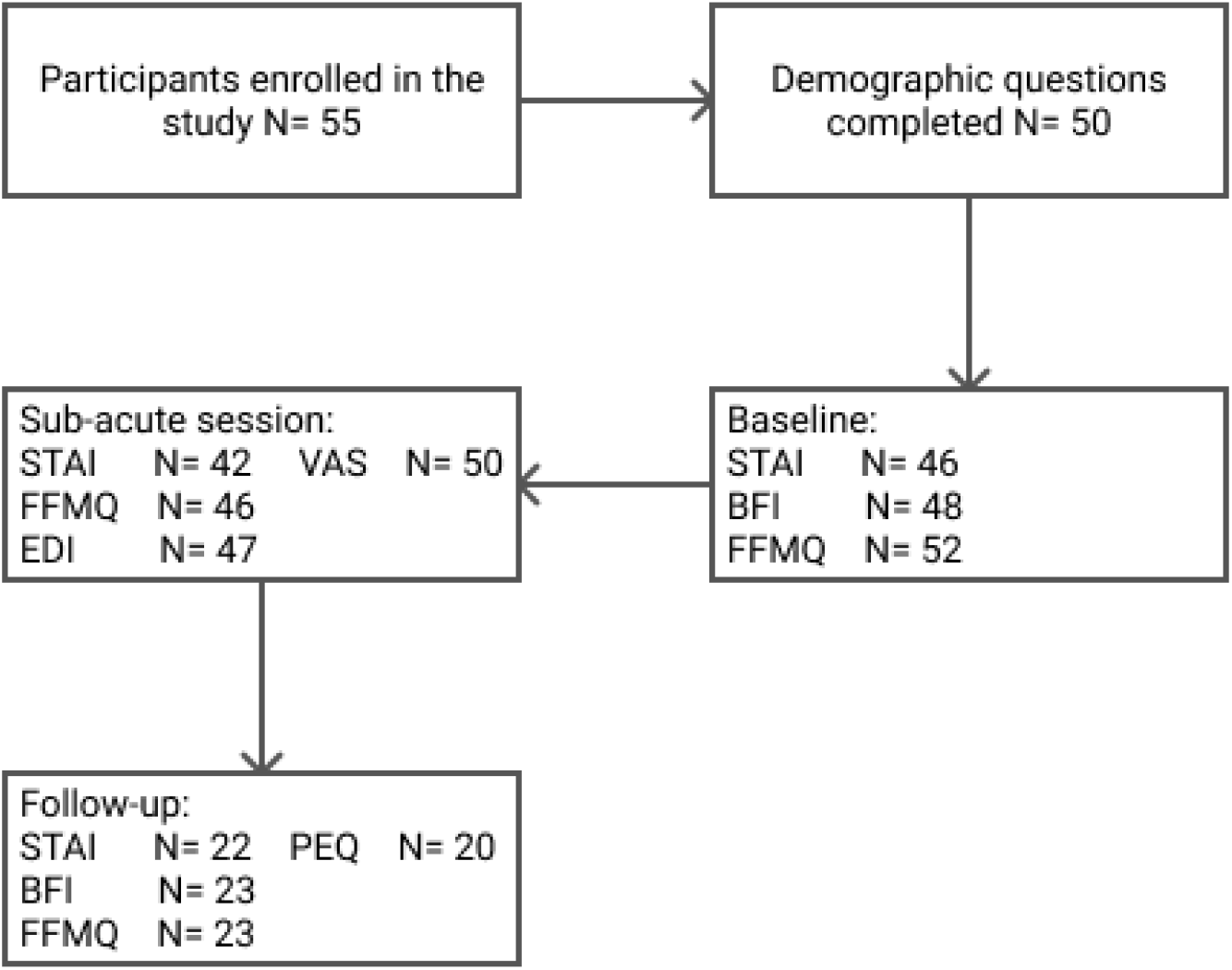
Flow-chart of participants’ enrollment and survey completion

The mean age of the entire group was 34.8 (SD = 8.9), ranging from 20 to 58 years. The highest completed levels of education were graduate school (41.8%), undergraduate school (41.8%), secondary school (7.3%), or undisclosed (9.1%). Most participants were from Europe (80%), while the rest were from North America (7.3%), Africa (3.6%), Central America (1.8%), Asia (1.8%), or undisclosed (5.4%). About half of the sample had previous experience with classical psychedelics (psilocybin= 52.7%; LSD= 40%; ayahuasca= 52.7%; DMT= 58%), MDMA (56%), and the majority had experience with cannabis (78%). For 69.1% of the participants, this was the first time taking a psychedelic in a retreat setting.

#### Psilocybin sample

The truffle sample (15 grams; Psilocybe Hollandia) contained 1.9 mg of psilocybin and 10.5 mg of psilocin. Participants ingested an average (SD) 34.2 (8.9) grams of truffles throughout the day. Once ingested, psilocybin is quickly metabolized to psilocin at a calculation factor of 0.719, resulting in a final (average) psilocin consumption of 27.1 mg.

### 3.4 State-Trait Anxiety Inventory (STAI)

Overall 46 (at baseline), 42 (at the morning after the ceremony), and 22 (at one week after the ceremony) participants completed all the parts of STAI and were included into analyses. The OLS multi-linear regression revealed a significant main effect of *Session* (F_2,107_= 14.10; *p*< 0.001) on scores capturing state anxiety. Compared to baseline, participants’ state anxiety reports were 6.4 points lower the morning after the psilocybin ceremony (*p*< 0.001; *d*= 0.77), and 6.7 points lower one week after the ceremony (*p*= 0.001; *d*= 0.87; Figure 2). *Session* was also found to have significant effects on estimates of trait anxiety (F_2,107_= 10.13; *p*= 0.002). Specifically, compared to baseline, self-rated trait anxiety was about 6 points lower the morning after the ceremony (*p*= 0.014; *d*= 0.52) and 8.6 points lower one week after the ceremony (*p*= 0.004; *d*= 0.77; Figure 3) on a 60-point range.

**Figure 2.**
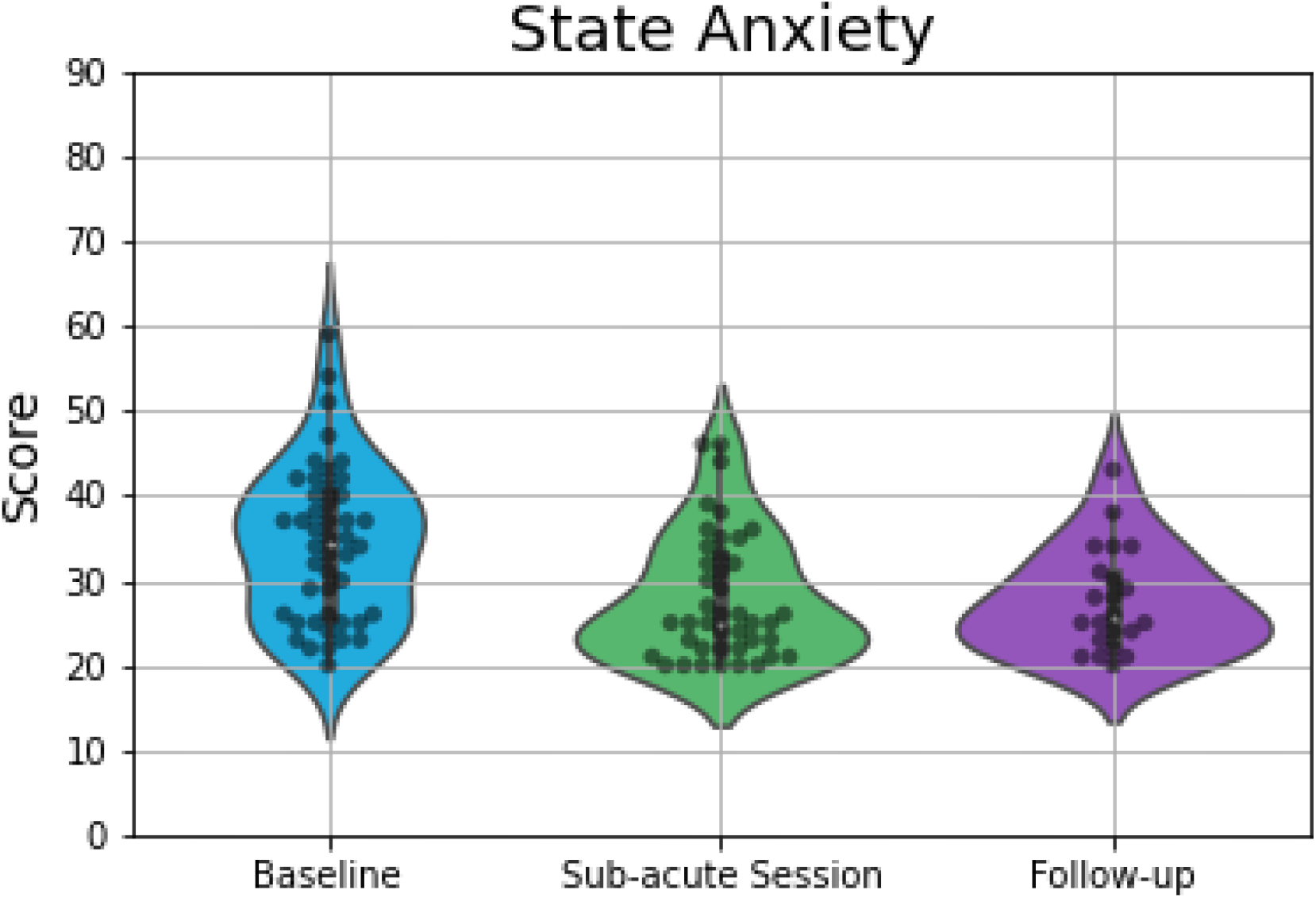
Violin plots displaying scores on measures of state anxiety, which demonstrated significant differences between baseline and the other two time points. The plot consists of median (SE) ratings of state anxiety before, 1-day after and 7-days after psilocybin truffles. The thick line indicates the interquartile range, whereas the white dot indicates the median. Each grey dot indicates a data point, whereas the density is scaled to the relative count across all bins. Wider sections of the violin plot represent a higher probability of observations of a given value.

**Figure 3.**
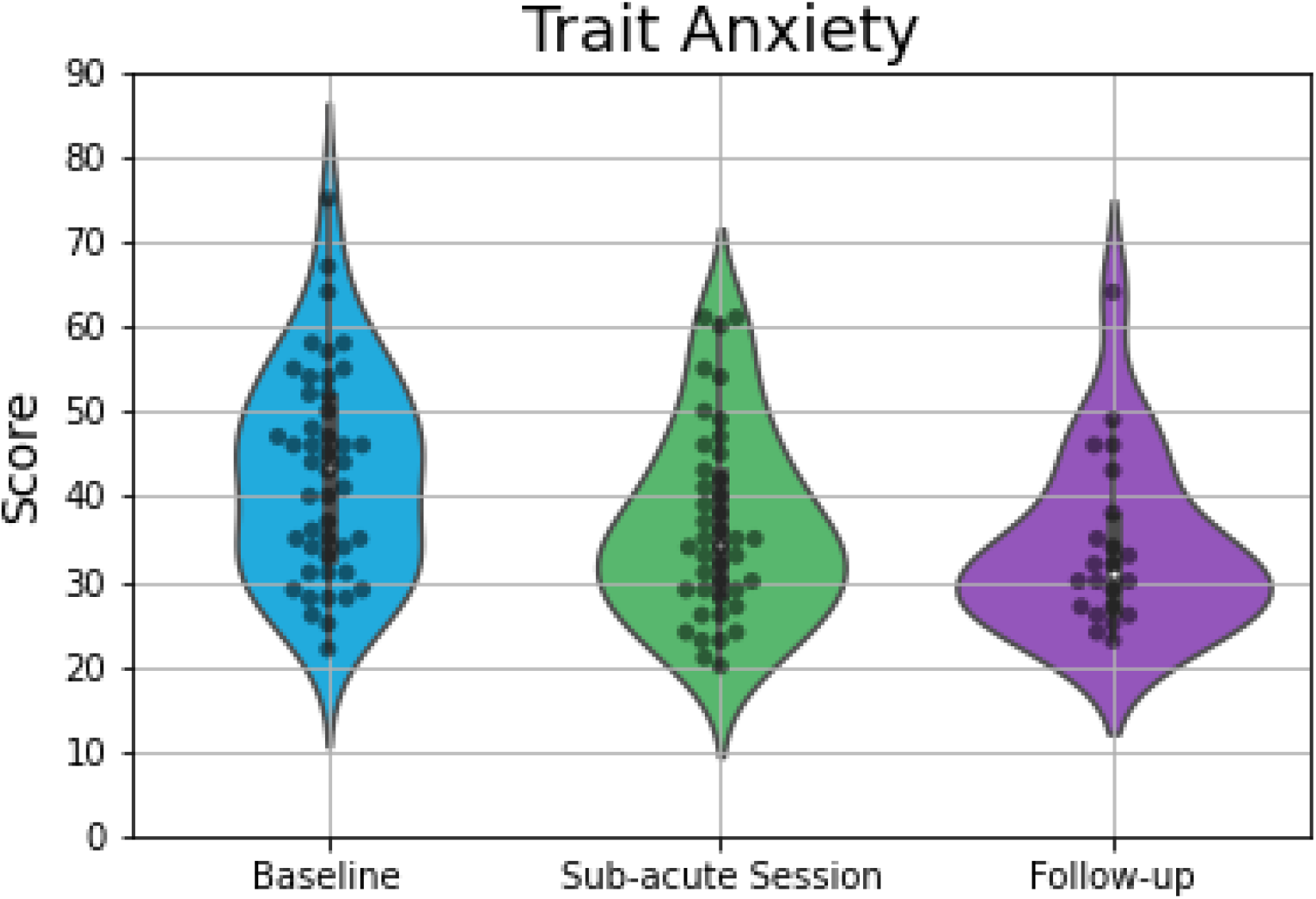
Violin plots displaying scores on measures of trait anxiety, which demonstrated significant differences between baseline and the other two time points. The plot consists of median (SE) ratings of trait anxiety before, 1-day after and 7-days after psilocybin truffles. The thick line indicates the interquartile range, whereas the white dot indicates the median. Each gray dot indicates a data point, whereas the density is scaled to the relative count across all bins. Wider sections of the violin plot represent a higher probability of observations of a given value.

### 3.5 Five Facet Mindfulness Questionnaire (FFMQ)

In total, 52 (at baseline), 46 (at the sub-acute session), and 23 (at the follow-up session) participants completed the FFMQ. The OLS multi-linear regression revealed a significant main effect of *Session* on the *Non-judge* dimension (F_2,118_= 4.37; *p*= 0.04) of the FFMQ. The contrasts analysis showed, in comparison to the baseline, a 0.5 increase in non-judgmental mindfulness capacities the morning after the ceremony (*p*< 0.39; *d*= 0.17) and 1.5 increase at the follow-up (*p*= 0.03; *d*= 0.51). There was no main effect of Session on the remaining, four facets (Table 1).

**Table 1.** Summary of regression and contrast analyses of the dependent measures

| Variable | F (*p*) | Session | Mean | SE | *p* | *d* |
| --- | --- | --- | --- | --- | --- | --- |
| State Anxiety | 14.1<br>($< 0.001$) | Baseline | 34.3 | 1.17 | | |
| | | Sub-acute | 27.9 | 1.7 | $< 0.001$ | 0.77 |
|  |  | Follow-up | 27.6 | 2.06 | 0.001 | 0.87 |
| Trait Anxiety |  | Baseline | 42.6 | 1.68 |  |  |
|  |  | Sub-acute | 36.55 | 2.42 | 0.014 | 0.52 |
|  |  | Follow-up | 34.05 | 2.94 | 0.004 | 0.77 |
| Observe<br>(FFMQ) | 0.11 (0.74) | Baseline | 10.27 | 0.43 |  |  |
|  |  | Sub-acute | 10.28 | 0.63 | 0.93 | 0.003 |
|  |  | Follow-up | 10.57 | 0.78 | 0.7 | 0.1 |
| Describe<br>(FFMQ) | 0.79 (0.38) | Baseline | 10.36 | 0.92 |  |  |
|  |  | Sub-acute | 12.24 | 1.34 | 0.17 | 0.25 |
|  |  | Follow-up | 11.35 | 1.66 | 0.56 | 0.34 |
| Acting with<br>awareness<br>(FFMQ) | 2.96 (0.09) | Baseline | 9.44 | 0.37 |  |  |
|  |  | Sub-acute | 10.39 | 0.54 | 0.08 | 0.36 |
|  |  | Follow-up | 10.39 | 0.66 | 0.16 | 0.36 |
| Non-judging<br>(FFMQ) | 4.37 (0.04) | Baseline | 11.36 | 0.4 |  |  |
|  |  | Sub-acute | 11.83 | 0.58 | 0.39 | 0.16 |
|  |  | Follow-up | 12.87 | 0.72 | 0.03 | 0.51 |
| Non-reacting<br>(FFMQ) | 0.12 (0.73) | Baseline | 9.63 | 0.36 |  |  |
|  |  | Sub-acute | 9.59 | 0.53 | 0.93 | 0.02 |
|  |  | Follow-up | 9.39 | 0.65 | 0.71 | 0.09 |
| Extraversion<br>(BFI) | 1.76 (0.19) | Baseline | 27.0 | 0.86 |  |  |
|  |  | Follow-up | 29.0 | 1.5 |  | 0.36 |
| Agreeableness<br>(BFI) | 8.87 (0.004) | Baseline | 33.96 | 0.7 |  |  |
|  |  | Follow-up | 37.74 | 1.27 |  | 0.78 |
| Conscientiousness<br>(BFI) | 3.6 (0.06) | Baseline | 28.8 | 0.97 |  |  |
|  |  | Follow-up | 32.0 | 1.7 |  | 0.47 |
| Neuroticism<br>(BFI) | 8.5 (0.005) | Baseline | 23.9 | 1.0 |  |  |
|  |  | Follow-up | 18.7 | 1.78 |  | 0.76 |
| Openness<br>(BFI) | 3.59 (0.06) | Baseline | 37.7 | 0.7 |  |  |
|  |  | Follow-up | 40.0 | 1.2 |  | 0.5 |

### 3.6 Big Five Inventory (BFI)

The BFI was completed by 48 and 23 participants at baseline and follow-up, respectively. The OLS multi-linear regression model revealed a significant main effect of *Session* on *Neuroticism* (F_1,69_= 8.46; *p*= 0.005) and *Agreeableness* (F_1,69_= 8.87; *p*= 0.004) of the BFI. The two traits showed an opposite pattern of changes over time. While, in comparison with baseline, participants scored 5.2 points lower on *Neuroticism* (*d*= 0.76) at the follow-up, they scored 3.8 points higher on *Agreeableness* (*d*= 0.78). There were no main effects of Session on the remaining three personality traits (Table 1).

### 3.7 Persisting Effects Questionnaire (PEQ)

In total, 20 participants filled in the PEQ 7 days after the ceremony. Mean (SE) of the positive and negative ratings assessing attitudes, mood, social effects, and behavior are presented in Table 2.

**Table 2.** Mean (SE) scores on outcome measures of the Persisting Effects Questionnaire

| Questionnaire subscales and single items | Mean (SE) |
| --- | --- |
| Attitudes about life |  |
| Positive | 59.30 (2.19) |
| Negative | 15.65 (0.67) |
| Attitudes about self |  |
| Positive | 45.30 (2.50) |
| Negative | 14.75 (0.71) |
| Mood changes |  |
| Positive | 37.55 (2.06) |
| Negative | 9.95 (0.41) |
| Social effects |  |
| Positive | 36.85 (2.38) |
| Negative | 10.30 (0.45) |
| Behavioural changes |  |
| Positive | 4.65 (0.28) |
| Negative | 1.15 (0.15) |

On the question, “how personally meaningful was the experience”, 2 (10%) rated it as the single most meaningful experience of their lives, whereas 10 (50%) and 2 (10%), rated it as among the 5 and 10 most meaningful experiences of their lives, respectively. Two participants (10%) rated it as similar to the meaningful experiences that occur on average once every 5 years, three (15%) stated it to be similar to meaningful experiences that occur once a year, and one (5.0%) stated it was similar to experiences that occur on average once a month.

On the question, “how spiritually significant was the experience”, 3 (15%) rated it as the most spiritually significant experience of their lives, whereas 8 (40%) and 3 (15%) rated it as among the 5 and 10 most spiritual experiences of their lives, respectively. Three participants (15%) rated it as similar to the spiritually meaningful experiences that occur on average once every year, 2 (10%) stated it was similar to spiritually meaningful experiences that occur once a month, and one (5.0%) stated it was no more spiritual than a routine, everyday experience.

In regards to how psychologically challenging the experience was, 1 (5%) rated it as the single most difficult or challenging experience of their lives, 6 (30%) rated it as among the 5 most challenging experiences of their life, and 1 (5%) rated it as among the top 10 most challenging experiences of their lives, followed by 3 (15%) stating it was similar to the challenging experiences that occur every 5 years, 3 (15%) who said occur every once a year, 4 (20%) who said occur once a month, 1 (10%) who said occur once a week, and 1 (10%) who stated it was no more psychologically challenging than a routine, everyday experience.

Finally, regarding the psychological insightfulness of the experience, 7 (35%) stated the experience to be the single most psychologically insightful experience of their life, 6 (30%) and 2 (10%) stated among the 5 and 10 most insightful experiences, respectively. Three (15%) stated that the experience was similar to psychologically insightful experiences that occur on average once every 5 years, one (5%) stated similar to experiences that occur on average once a month, and one (5%) stated the experience was no different from every psychologically insightful experience.

### 3.8 Ego Dissolution Inventory (EDI)

The EDI was completed by 47 participants the morning after the ceremony. The mean (SD) of participants who filled in the Ego Dissolution Inventory was 59.7 (28.3), with ratings varying between 3 (minimal reported score) and 100 (maximal reported score). The kernel density estimate of the probability density function of the individual EDI scores is presented in Figure 4.

**Figure 4.**
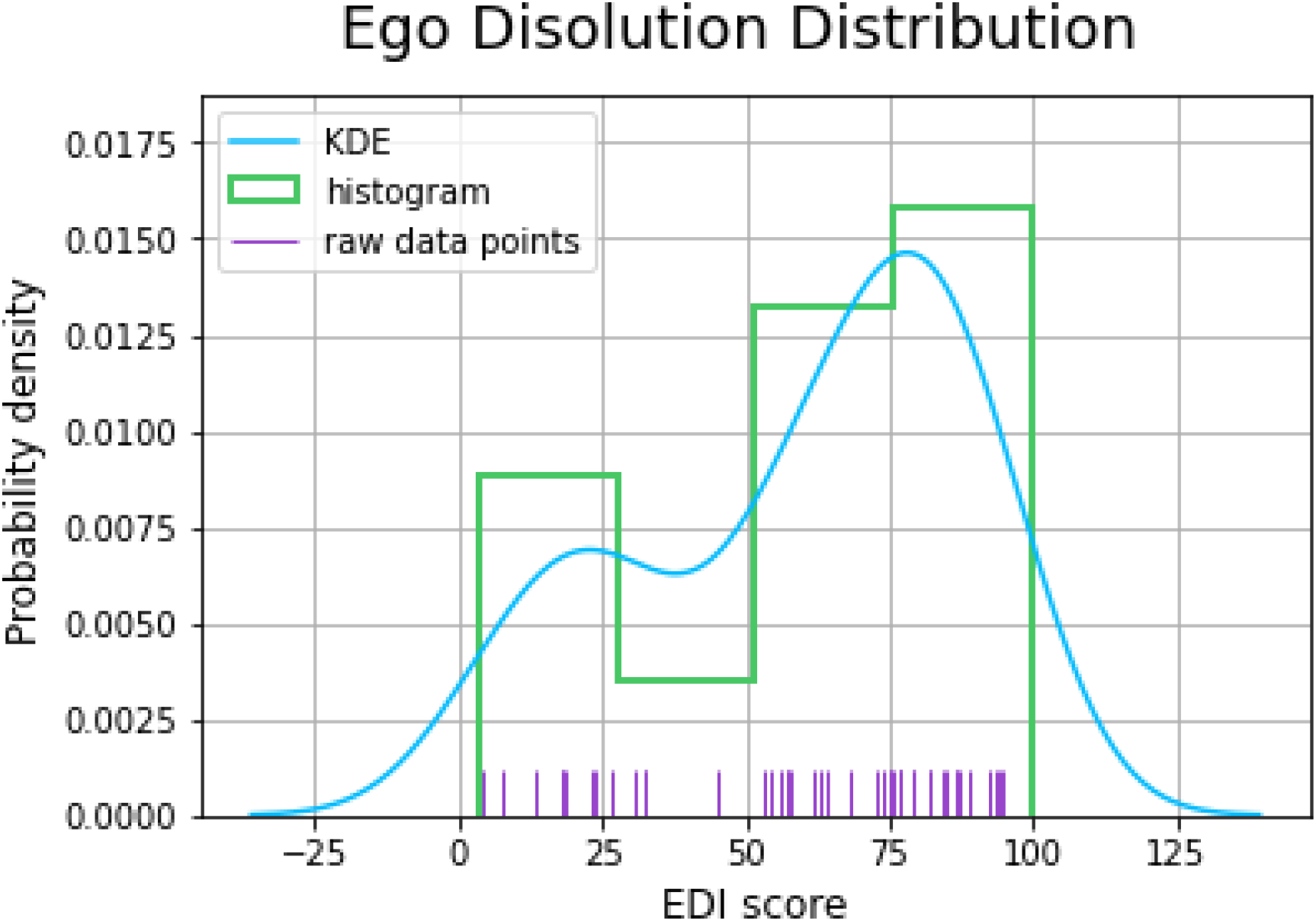
Probability distribution plot of the experience of ego dissolution as assessed by the EDI morning after the psilocybin ceremony. The plot depicted kernel density estimation (KDE), histogram, and raw data points.

### 3.9 Hypothesis-driven correlations

A canonical correlation analysis was conducted using the three psychological variables which demonstrated significant effects as predictors (follow-up scores on non-judge dimension of FFMQ, follow-up neuroticism scores, and EDI scores) of the four anxiety outcome measures: sub-acute and follow-up scores on state and trait anxiety measures. The analysis yielded three functions with squared canonical correlations (*R*_c_^2^) of 0.891, 0.324, and 0.096 for each successive function. The full model across all functions was statistically significant *F*_(12,18.81)_=2.81, *p*=0.022), explaining 93.4% of the variance. From this model, the first two functions were considered noteworthy in the context of this study, explaining 89.1% and 32.4% of the variance, respectively.

Table 3 presents the standardized canonical function coefficients, the structure coefficients (*r*_s_), and the squared structure coefficients (*r*_s_^2^) for each function, as well as the communalities (*h*^2^) across the functions for each variable (for an in-depth explanation and interpretation of canonical correlations and associated terminology, the reader is referred to Sherry and Henson 2005). Function 1 indicated that the dominant contributors were sub-acute and long-term changes in trait anxiety, whereas the dominant predictor was the change in neuroticism, and the secondary predictor was changes in non-judgement. Overall, this function suggests that the strongest predictor of changes in *trait* anxiety were changes in neuroticism.

**Table 3.** Canonical solution for psychological variables predicting state and trait anxiety for Functions 1 and 2. *r*_*s*_ greater than |.45| and *h*^*2*^ greater than 45% are underlined and deemed valuable contributors.

| Variable | Function 1 |  |  | Function 2 |  |  |  |
| --- | --- | --- | --- | --- | --- | --- | --- |
| | Coef | $r_s$ | $r_s^2$ (%) | Coef | $r_s$ | $r_s^2$ (%) | $h^2$ (%) |
| State anxiety (subacute) | -.097 | <u>-.494</u> | 24.40 | -.353 | <u>-.528</u> | 27.88 | <u>52.28</u> |
| Trait anxiety (subacute) | -.312 | <u>-.819</u> | 67.08 | 1.04 | .065 | 0.42 | <u>67.50</u> |
| State anxiety (followup) | .331 | <u>-.526</u> | 27.67 | -.997 | <u>-.712</u> | 50.69 | <u>78.36</u> |
| Trait anxiety (followup) | -.904 | <u>-.963</u> | 92.74 | -.156 | -.227 | 5.15 | <u>97.89</u> |
| $R_c^2$ | | | 89.15 | | | 32.43 | |
| Non-judging (followup) | -.221 | <u>.547</u> | 29.92 | -.267 | .303 | 9.18 | 39.10 |
| Neuroticism (followup) | -1.189 | <u>-.974</u> | 92.23 | 0.136 | -.157 | 2.46 | <u>94.69</u> |
| EDI | -.126 | .246 | 6.05 | 1.160 | <u>0.949</u> | 90.06 | <u>96.11</u> |

Coef = standardized canonical function coefficients; $r_s$ = structure coefficients; $r_s^2$ = squared structure coefficient; $h^2$ = communality coefficient; $R_c^2$ = squared canonical coefficient

Function 2 indicated that the dominant contributors were sub-acute and long-term changes in state anxiety, with the dominant predictor of ratings of EDI. Overall, this function suggests that the strongest predictor of changes in *state* anxiety was higher ratings in ego dissolution.

## 4. Discussion

The impact of anxiety disorders on people’s lives and the treatment challenges are well-known within the field of mental health (Brown and Barlow 1995; Brown et al. 1996; World Health Organization 2017; Otto et al. 2010; Stein et al. 2017). The current study investigated the effects of a single administration of psilocybin on ratings of state and trait anxiety across retreat attendees, when taken in a supportive naturalistic setting. We further explored hypothesized psychological processes which may predict reductions in anxiety. Compared to baseline measures, we observed medium to large reductions in state anxiety ratings, and medium decreases in trait anxiety ratings, which persisted over a one week period post the ingestion of psilocybin. Additionally, we found enhancements in self-rated mindfulness capacities and alterations in personality traits at one week post ceremony. Regarding the post-retreat changes in anxiety, analyses showed that ratings of neuroticism and ego dissolution were the most strongly correlated with reductions in trait and state anxiety, respectively.

The present study’s findings are in line with results from historical (clinical) studies, which suggest reductions of anxiety symptoms after a combination of psychedelic drug administration and psychological therapy. From this work, four paradigms (and success rates) emerged in which psychedelic psychotherapy was used: 1. a single LSD session, provided after an intensive psychoanalytic session (56%); 2. repeated LSD sessions used in combination with individual psychotherapy (56%); 3. combination of repeated LSD sessions with individual psychotherapy completed by additional group therapy (62.5%); 4. sessions with psychotomimetics only applied in the course of group therapy (40%) (Mascher 1967). Similarly, recent clinical studies have found reductions of anxiety in patients with life-threatening illness (Gasser et al. 2014; Griffiths et al. 2016; Ross et al. 2016) and comorbid treatment-resistant depression (Carhart-Harris et al. 2018; Carhart-Harris et al. 2016; Osório et al. 2015). Trials with healthy volunteers in the laboratory, however, have shown mixed results. Whereas one study showed reductions in state anxiety one week post administration, and reductions in trait anxiety one month post-administration, of a high dose of psilocybin (25 mg/70 kg), another study found no changes in trait anxiety one and 12 months after a high dose of LSD (200 ug) (Barrett et al. 2020; Schmid and Liechti 2018). Finally, previous naturalistic work with individuals reporting heterogenous mental health status also found reductions in anxiety after participation in a 5-MeO-DMT ceremony (Uthaug et al. 2019). That said, a more recent placebo-controlled naturalistic study found that state anxiety ratings decreased 24 hours after volunteers ingested both the placebo and the psychedelic substance, ayahuasca (Uthaug et al. 2021). Taken together, the majority of modern studies present some evidence for psychedelic-induced reductions in anxiety symptoms across diverse populations and study designs. Studies suggest that not only the momentary anxiety estimates (state anxiety), but also the more stable personality-inherent features of anxiety proneness (trait anxiety) showed significant decreases. However, given some of the confounding findings (Schmid and Liechti 2018; Uthaug et al. 2021), future research is needed to further explore the role of extra pharmacological factors (e.g., setting, dose, design) on the psilocybin-anxiety interaction.

In line with previous studies (Erritzoe et al. 2018; Johnstad 2021; Kiraga et al. 2021), we detected long-term decreases in trait neuroticism, which has been shown to be strongly associated with the acuity and comorbidity levels of a range of mental disorders (Bienvenu et al. 2004; Cuijpers et al. 2005). Interestingly, levels of neuroticism have been found to correlate positively with frontolimbic serotonin 5-HT_2A_ receptor binding, the latter being the main target of psychedelic drugs (Frokjaer et al. 2008; Frokjaer et al. 2010), suggesting direct 5-HT_2A_ agonism may be the biological basis by which these substances alter neuroticism. Findings from the current and previous studies suggest that a psychedelic-assisted intervention can reduce levels of neuroticism, which are related to reductions in anxiety ratings in our study, and could be related to reductions of symptoms of comorbid conditions such as depression or post-traumatic stress disorder as seen in other studies. We also found post-retreat increases in trait agreeableness, which is in line with a previous naturalistic work done by Weiss et al. (2021). Other studies have shown either no changes in agreeableness post-psilocybin experience in depressed patients (Erritzoe et al. 2018) or higher agreeableness scores among psychedelics users when compared to the general population (Johnstad 2021). While the psychedelics’ effects on neuroticism are more robust and more frequently reported, the emerging effects on agreeableness are novel and their relevance to therapeutic outcomes in patient populations has yet to be determined.

Lastly, the present study aimed to further explore hypothesis-driven correlational relationships between changes in mindfulness and neuroticism and changes in anxiety. Although there was an increase in mindfulness capacities following intake of psilocybin, there were no strong associations between changes in mindfulness capacities and anxiety ratings. In regards to neuroticism, in line with theoretical conceptualizations of neuroticism and trait anxiety as stable, personality-dependent dispositions, we found that sub-acute and long-term reductions in trait anxiety were most strongly correlated with reductions in neuroticism. Namely, change scores in neuroticism and trait anxiety shared about 89% of the variance variability, pointing out a strong interdependency between the two variables. Additionally, we also found that the strongest correlator of sub-acute and long-term changes in state anxiety were ratings of ego dissolution, sharing about 32% of the variance. Specifically, the higher the rating of ego dissolution during the acute experience, the larger the decrease in state anxiety scores both 24 hours and one week after the experience. A recent review of twenty studies assessing the clinical response to psychedelics in patients with a range of psychological disorders concluded that the main predictive factor of a response to a psychedelic is the intensity of the acute psychedelic experience (Romeo et al. 2021). That said, it has yet to be systematically assessed whether such a “peak” experience is necessary for long-term outcomes (Olson 2021), or whether the subjective experiences elicited by psychedelic substances are merely epiphenomena of the underlying neurobiological mechanisms, the latter which are conveying any beneficial effects. Additionally, it is possible that other psychological components play important roles in mediating long-term outcomes of psychedelic experience, for instance *insight/breakthrough* or *catharsis, suggestibility*, and *reliving of trauma* have been suggested as important factors determining the psychedelic experience (Belser et al. 2017; Eisner and Cohen 1958; Frederking 1955; Gasser et al. 2014; Richards 1978; Russ and Elliott 2017; Watts et al. 2017). Therefore, we should remain cautious when developing and interpreting specific theoretical frameworks of psychedelics’ (psychological) mechanisms of action and focus future efforts on testing these frameworks in the context of experimental studies.

The objectives were tested using a naturalistic, observational design, with attendees of psilocybin ceremonies. Although, this sort of setting has high rates of ecological validity, as it closely resembles a typical environment associated with psychedelic usage (Frecska et al. 2016; Hartogsohn 2016; Lawn et al. 2017; Sapoznikow et al. 2019), it also comes with a range of confounding variables. In a naturalistic setting there is no control over the ceremonial setting and substance administration (Uthaug et al. 2021). To have some indication of the amount of truffles individuals took, we asked for a sample that was analyzed afterward and we asked how much truffles participants consumed. This allows comparisons between findings of our study and that of controlled studies. Another point is that longitudinal studies traditionally come with high dropout rates (Chatfield et al. 2005; Young et al.2006); our study was not different in that sense. The concern of such dropouts is that they could create problematic biases in the data, with for example only those individuals experiencing benefits from the experience being motivated to continue responding to the questions. A recent study investigating potential determinants of study attrition in web-based prospective studies on psychedelic use identified that baseline predictors of attrition (i.e., age, educational levels, and personality traits) were consistent with those reported in longitudinal studies in other scientific disciplines, suggesting their transdisciplinary relevance (Hübner et al. 2021). Moreover, they did not find an association between attrition and psychedelic advocacy or negative drug experiences, advocating against the concerns about problematic biases in these and related data.

## Conclusion

Taken together, the present study suggests persisting reductions in anxiety symptoms and trait neuroticism, and increases in mindfulness capacities, after ingestion of psilocybin in a supportive group setting. Given the important role of set and setting on the outcome of psychedelic experience (Hartogsohn 2016; Sapoznikow et al. 2019), future studies should systematically evaluate the contribution of these extra-pharmacological factors to the treatment outcome, in order to optimize therapeutic strategies. Additionally, naturalistic retreat studies such as this, as well as modern clinical trials (Anderson et al. 2020), are beginning to demonstrate feasibility, safety, and efficacy of group sessions with psychedelics, across different disorders. As it is likely that psychedelic-assisted psychotherapy will be costly and time-consuming for both the patient and the practitioners, future studies exploring the feasibility of group therapy could potentially substantially lower treatment costs, resulting in a more financially accessible treatment option.

## Data Availability

All data produced in the present study are available upon reasonable request to the authors

## Acknowledgements

The authors would like to acknowledge and thank Stefana Bosse and the Psychedelic Society UK for their collaboration and permission to collect data at their experience weekends. They would like to thank Hannes Kettner and Beatrice Da Rios for helping with data collection. They would also like to thank all participants for their time and effort.

